# ANALYSIS OF VIRAL LOAD IN COLOSTRUM OF PUERPERAL WOMEN LIVING WITH HIV USING ANTIRETROVIRAL THERAPY (ART)

**DOI:** 10.1101/2024.08.04.24311472

**Authors:** Raphaela Barbosa Gonçalves Souza, Luiz Claudio Pereira Ribeiro, Renata Dias Reis, Bárbara Motta, Julia Rodrigues Carvalho Ancora Luz, Thais Moraes Araújo, Julia Freitas Fernandes Santos, Carolina Romão Azevedo, Julia Monteiro Jacarandá, Sandy Borges de Aguiar, Isabella Navarro Diaz Horta, Gustavo Mourão Rodrigues, Barbara Rodrigues Geraldino, Regina Rocco, Rafael Braga Gonçalves

## Abstract

HIV transmission occurs during pregnancy, childbirth, and breastfeeding, and can be mitigated through measures such as prenatal HIV screening and antiretroviral therapy (ART). Health institutions are gradually releasing it if a woman has an undetectable viral load in her blood. This study aims to determine the viral load of HIV in colostrum from postpartum women using qRT-PCR, following participant selection based on informed consent criteria. The research includes monitoring participants’ prenatal routine, collecting, storing, and processing colostrum, as well as automated RNA extraction and viral RNA quantification via qRT-PCR. Thirteen participants were recruited, 13 samples of colostrum were subsequently collected, meeting eligibility criteria and signing informed consent. These samples were processed and analyzed by qRT-PCR. Six of nine samples analyzed were undetectable, and three were below the detection limit. We observed that patients with undetectable colostrum had been on ART before conception, while those with colostrum samples detectable had a detectable serum viral load at some point during pregnancy. The immunological, biochemical, and socioeconomic impact of non-breastfeeding on maternal-child health is significant, and analyzing the transmissibility potential of colostrum raises questions about possible breastfeeding with reduced risks of HIV transmission. We think that achieving undetectable or below detection limit levels in colostrum of women on ART is feasible, but further research is needed on the condition of breast milk from women living with HIV under current antiretroviral therapies used in Brazil.

## 1. Introduction

In recent years, the epidemiological paradigm “Undetectable=Untransmittable” (U=U) has been established based on clinical evidence. This concept summarizes that individuals with HIV, who achieve and sustain an undetectable viral load through a consistent antiretroviral therapy regimen cannot transmit the virus. This advance represents a significant shift in understanding HIV transmission and has important implications for public health strategies (Hui, 2023).

In Brazil, from 2000 to 2023, 158.429 pregnant women living with HIV (WLH) were reported, 7.943 of which occurred in 2022, with a detection rate of 3.1 per 1,000 live births (Brazil, 2023). Despite the risks of HIV transmission, breastfeeding is a practice embraced in several countries, justifying that an undetectable viral load reduces infection rates to extremely low levels or even nullifies this risk (Penazzato et al., 2023).

The World Health Organization (WHO) since 2010, recommends that infants from WLH with undetectable viral load should exclusively breastfeed until six months of age, since the first month of life is crucial for infant survival. Just in 2022, approximately 2.3 million newborns died, highlighting the persistent challenges in reducing neonatal mortality (WHO, 2010). Despite a significant drop in mortality rates since 2000, even in 2022, almost half (47%) of all deaths of children under five years occurred in the first 28 days of life, underscoring the importance of effective interventions in the neonatal period (WHO, 2024).

Breastfeeding is associated with a reduced risk of developing certain chronic conditions such as type 2 diabetes and cardiovascular diseases in infants. Additionally, the benefits of exclusive breastfeeding in reducing the risk of gastrointestinal and respiratory infections in infants are well documented (Tschiderer et al., 2022). Breastfeeding is also related to a lower risk of postpartum depression and the strengthening of the emotional bond (Alimi et al., 2022; Eccles et al., 2022).

Strategies such as HIV screening during pregnancy, use of antiretroviral therapy (ART), and non-breastfeeding almost prevent vertical transmission of HIV in Brazil (SBP, 2023). Although breastfeeding is associated with an additional risk of transmission, this is significantly reduced when the standard treatment protocol is rigorously followed (Bamford et al., 2024). In low-income countries, children die due to socio economic difficulties breastfeeding is an effective recommendation since the weighted risk of children dying from other factors is extremely high (Engelhart et al., 2022).

In Brazil, the Guidelines for pregnant and postpartum women living with HIV recommend replacing breastfeeding with formula feeding as a preventive measure against vertical transmission of HIV. Based on WHO and NIH recommendations, local studies are crucial to guide health practices and proper public policies (WHO, 2016).

Although the possible risk associated with HIV transmission through breastfeeding is known, WHO advises that these women breastfeeding exclusively during the first six months and continue up to 12 months, provided they are on effective ART without clinical, immunological, or virological failures. Additionally, WHO advises that, with adequate treatment, women with HIV can extend breastfeeding up to 24 months or more, thus aligning with the recommended practices for the general population (WHO, 2016).

Recently, high-income countries are counseling breastfeeding in t women with an undetectable viral load. In the United States, policies support patient-centered shared decision-making HIV parents. (NIH guideline) (Spat, 2023).

The objective of this study was to quantify the viral load in the colostrum of postpartum women attended at the Gaffrée and Guinle University Hospital (HUGG), a reference in high-risk pregnancy (HIG), using the Real-Time PCR (qRT-PCR) technique.

## 2. Material and Methods

### 2.1. Ethical Aspects and Protection of Research Participants

The research protocol for the current study was developed in accordance with Resolution 466 of December 12, 2012, of the National Health Council on research involving human beings. This study was submitted for evaluation by the Research Ethics Committee on human beings at the Gaffrée and Guinle University Hospital (HUGG) of the Federal University of the State of Rio de Janeiro (UNIRIO), registered, and approved under the number CAAE: 61885722.1.0000.5258.

### 2.2. Inclusion and Exclusion Criteria

The study recruited pregnant women aged between 18 and 45 years, with a confirmed diagnosis of human immunodeficiency virus (HIV) infection, evidenced by a positive rapid test and detectable viral load at the time of diagnosis. Participants were required to be available for follow-up throughout the study period and have an undetectable viral load at 34 weeks of gestation. Additionally, they needed to have negative results for HBV, HCV, and HTLV tests in the last trimester of pregnancy, be on continuous antiretroviral therapy (ART) during pregnancy and be able to understand and sign the Free and Informed Consent Form (ICF). Exclusion criteria for the study participants included pregnant women with other immunosuppressive or autoimmune pathologies, use of immunosuppressive medications besides ART, patients with irregular or interrupted ART use, and those with a detectable viral load at 34 weeks of gestation or without a viral load result for that gestational age. Pregnant women with positive or no results in serological laboratory tests (HBV, HCV, and HTLV) in the last trimester of pregnancy or those who had received blood products in 60 days before sample collection were also excluded.

### 2.3. Sample Size Calculation

The sample size for this study was calculated using Cochran’s method, resulting in the participation of 29 pregnant women. This sample was chosen to ensure statistical precision and representativeness, with a confidence level of 95% and a margin of error of 10%, considering a population of 50 HIV-positive pregnant women attended annually. Patients were screened from the obstetrics and infectious diseases services from Gaffrée and Guinle University Hospital (HUGG).

### 2.4. Laboratory Analysis

Colostrum samples were initially centrifuged for 5 minutes at 9,600 g. After this centrifugation, the samples were refrigerated at -20ºC after discarding the supernatant. Then, samples were centrifuged again for 5 minutes at 9.600 g. The resulting liquid was directed to the viral extraction process. The quantification of the HIV-1 viral load was performed using the Cobas® HIV-1 Test with the automated Cobas® 4800 system (Roche).

## 3. Results

Twenty-seven pregnant women were recruited between March 2023 and July 2024. Seventeen delivered and ten are still pregnant. Sixteen participants initially selected, two did not achieve lactation. Additionally, two other patients chose not to participate, as they did not feel well enough to perform the milk expression (Figure 1).

**Figure 1:**
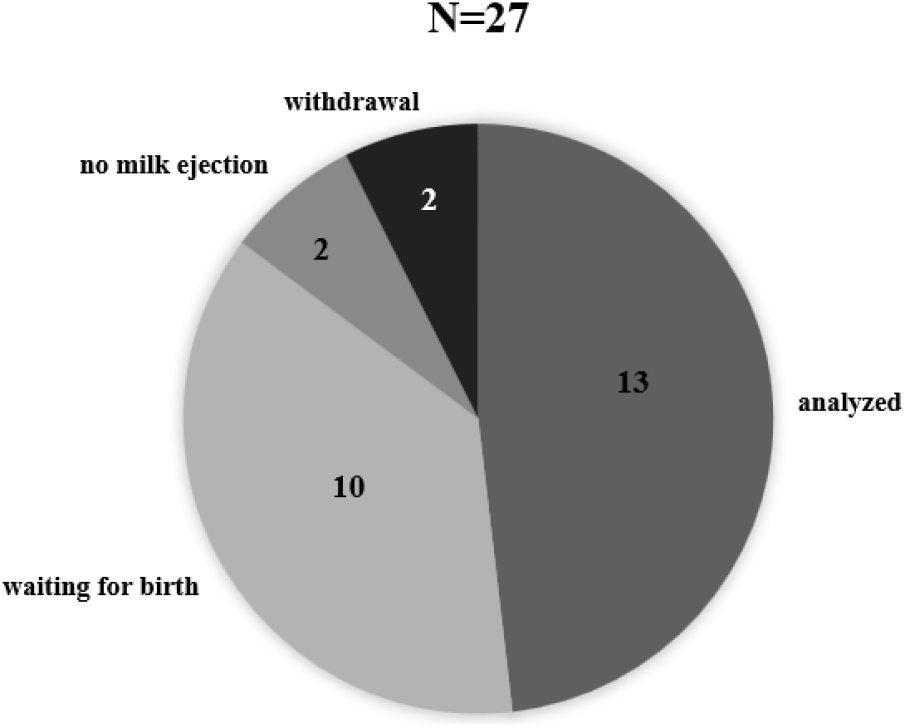
Twenty-seven pregnant women living with HIV were selected over a period of 16 months according to the study’s inclusion/exclusion criteria. The data in the graph represents this universe of participants.

Of the 13 samples analyzed, six were undetectable, four were below the detection limit, and three had detectable viral loads. It was observed that patients with undetectable colostrum had either a history of prior ART use before pregnancy or started ART in the first trimester of pregnancy, while patients with samples below the detection limit had positive viral loads at some point during pregnancy. Only one patient who used ART before pregnancy had a viral load below the detection limit and had a history of irregular medication use before pregnancy. Among the participants with detectable results, one had a history of irregular medication dispensation, and two patients had detectable viral loads despite regular medication use.

Regarding the treatment regimen, one patient was on the lamivudine (3TC), tenofovir (TDF), and atazanavir (ATV)/ritonavir (RTV) regimen, while the others were on the standard tenofovir, lamivudine, and dolutegravir (DTG) regimen (TABLE 1).

**Table 1:** Viral load on colostrum samples.

| Sample | Year of diagnosis | Start of ART | ART protocol used* | qRT-PCR result | Obs. |
| --- | --- | --- | --- | --- | --- |
| 00-1 | 2021 | during pregnancy | TDF+3TC+DTG | < min limit | - |
| 00-2 | 2021 | during pregnancy | TDF+3TC+DTG | < min limit | - |
| 00-3 | 2021 | pre-pregnancy | TDF+3TC+DTG | < min limit | irregular pre-pregnancy ART |
| 00-4 | 2007 | during pregnancy | TDF+3TC+DTG | undetectable | - |
| 00-5 | 2023 | during pregnancy | TDF+3TC+DTG | undetectable | - |
| 00-6 | 2013 | pre-pregnancy | TDF+3TC+ATV/RTV | undetectable | - |
| 00-7 | 2020 | during pregnancy | TDF+3TC+DTG | < min limit | - |
| 00-8 | 2023 | during pregnancy | TDF+3TC+DTG | undetectable | - |
| 00-9 | 2023 | during pregnancy | TDF+3TC+DTG | undetectable | - |
| 0-10 | 2023 | during pregnancy | TDF+3TC+DTG | 20.3 copies/mL | - |
| 0-11 | 2010 | pre-pregnancy | TDF+3TC+DTG | 106 copies/mL | start ART at 2016 |
| 0-12 | 2021 | pre-pregnancy | TDF+3TC+DTG | 20.5 copies/mL | - |
| 0-13 | 2023 | during pregnancy | TDF+3TC+DTG | undetectable | - |

\*TDF (tenofovir); 3TC (lamivudine); DTG (dolutegravir); ATV (atazanavir); r (ritonavir).

## 4. Discussion

Breastfeeding could be a HIV transmission route, and currently, data from the literature supports an absolute contraindication of this practice for women living with the virus (Montenegro e Rezende-Filho, 2017; Dunn et al., 1992). Despite the WHO’s defense of the right to choose breastfeed regardless of viral status, different health organizations still recommend the replacement of human milk from women living with HIV for artificial formulas to eliminate the risk of postnatal transmission (Ministério da Saúde, 2010; WHO, 2016; Gilleece et al., 2019)

Because the immunological, biochemical, and socioeconomic impact of not breastfeeding on the maternal-infant dyad is significant, it is necessary to seek new information about the security of breast milk from these women.

Colostrum is the milk produced in small quantities after the first few days postpartum and is rich in immunological components such as Immunoglobulin A (IgA), lactoferrin, leukocytes, and developmental factors like epidermal growth factor (EGF) (Ballard and Morrow, 2013; Menchetti L, 2016). IgA is correlated with the first line of mucosal immune defense, with higher concentration levels in colostrum (12 mg/mL) compared to mature milk (1 mg/mL), making colostrum a valuable immunological protection for the newborn (Van de Perre, 2003).

Among the functions of IgA are intracellular neutralization, viral excretion, and immunological exclusion. In intracellular neutralization, IgA undergoes transcytosis from the basolateral epithelial surface and targets endosomes containing viral particles for neutralization. Viral excretion involves viruses trapped in immune complexes that can undergo transcytosis on the basolateral surface and are excreted into the lumen. Finally, immunological exclusion occurs when immunoglobulin adheres to microorganisms, inhibiting their interaction with mucosal surfaces. IgA can also agglutinate bacteria and interfere with their motility by interacting with their flagella (Van de Perre, 2003; Rio-Aige, 2021). All these mechanisms contribute to protection in the early months of life and are exclusively acquired through breastfeeding.

Besides the immunological aspects involved, there are differences between the bioactive components of breast milk and infant formulas: formulas have standardized compositions, whereas breast milk has a dynamic composition that varies according to the timing of feeding during the day, the course of lactation, and even among different maternal populations (Ballard and Morrow, 2013). Among the bioactives in breast milk are epidermal growth factor (EGF), involved in intestinal maturation and repair; erythropoietin, which contributes to intestinal development and anemia prevention; calcitonin and somatostatin, which are growth-regulating hormones; adiponectin, involved in metabolism regulation and body growth; among others (Ballard and Morrow, 2013; Menchetti L, 2016). Thus, the components of breast milk are broadly beneficial and act in various systems of the child.

Moreover, the inhibition of this maternal process can increase the risk of gestational and postpartum depression (Llewellyn, Stowe and Nemeroff, 1997; Pereira et al., 2023). It has been observed that women who wished to breastfeed but could not do so experienced an increase in depression cases, and this clinical condition can have future repercussions. Possible outcomes of gestational depression include inadequate care and prenatal follow-up, insufficient nutrition, increased premature births, low birth weight, and preeclampsia (Waitt et al., 2018).

Regarding postpartum depression, there is an increased risk of future depression, with rates ranging from 50% to 62%. Additionally, this condition can lead to decreased productivity and the need for more extensive health care. Besides maternal harm, the negative effects of maternal depression can be observed in the cognitive, social, and physical development of the baby, such as anger, passivity, withdrawal, inappropriate attention and arousal mechanisms, which lead to other problems at different stages of the child’s life (Waitt et al., 2018; SPB, 2023)

The analysis of differences between breast milk and infant formulas goes beyond physiological aspects and involves socio economic issues. Several studies have shown that the HIV-positive population is predominantly from lower socioeconomic classes (Medeiros et al., 2017) and current epidemiological data illustrate the trend that HIV/AIDS case notifications are higher in conditions of social exclusion, poverty, and low educational levels in Brazil (Ministério da Saúde, 2007; 2021). The Public Health System is the only one providing treatment for HIV infection and supplying formulas for mothers living with HIV, but access to potable water and means to maintain this type of feeding are not provided by SUS, making access to adequate nutrition for their children a problem. The use of non-potable water and the lack of sterilization methods for feeding bottles can lead to a higher number of gastrointestinal infections and consequently increased healthcare needs (Bansaccal et al., 2020).

Besides the psychosocial implications of breastfeeding, it is necessary to address the laboratory results presented in this study in contrast to current guidelines and previous literature on postnatal vertical transmission. With the advent of new drugs in the last century and the possibility of achieving undetectable viral status and thus, untransmittable via sexual routes (U=U), it has been questioned whether this applies to breastfeeding (WHO, 2010). According to the WHO, in a population on continuous ART, the risk of maternal-fetal transmission is less than 1% in women who do not breastfeed, and this rate can reach 2% to 5% in women who choose to breastfeed (Freeman-Romilly et al., 2019).

Numerous studies on vertical transmission have compared outcomes in developed and developing countries. In 2020, a randomized study with 231 women living with HIV on ART was conducted in India and Sub-Saharan Africa. In these regions, breastfeeding is recommended due to low public health support, economic and nutritional vulnerability, where access to potable water for formula preparation and access to industrialized products is scarce. It was observed that with 6 months of breastfeeding, the risk of transmission was 0.3% (95% CI OR 0.1 to 0.8%), and with 12 months of breastfeeding, the risk increased to 0.7% (95% CI OR 0.3 to 1.4%) (SPB, 2018). It is concluded that the result with six months of breastfeeding, the time recommended for exclusive breastfeeding by the Brazilian Society of Pediatrics, showed statistical significance with low transmission rates (Bansaccal et al., 2020).

In developed countries, there is a lack of clinical studies due to access to artificial feeding (SPB, 2018). There have been reported cases of women who breastfed despite health recommendations, such as in Belgium, where two women chose to breastfeed their children. Both had undetectable viral loads when they became pregnant and remained so during pregnancy and breastfeeding. Maternal, infant, and milk viral loads were collected and analyzed monthly, with the first case having a breastfeeding duration of four months and the second five months. All results were negative, and at 18 months of age, both babies were declared HIV-negative (Weiss et. al., 2022).

In Germany, a cohort study was developed in which vertical transmission from women living with HIV who chose to breastfeed was observed over 5 years. In this study, babies received zidovudine as prophylaxis until the eighth week of life. There were 30 women, of whom 5 had positive viral loads during breastfeeding. No cases of vertical transmission were confirmed (Fogel et al., 2013).

According to the British HIV Association, women who choose to breastfeed should receive proper recommendations about the risks of such actions and be informed about the “safety triangle.” This triangle presents three main aspects for the safest possible breastfeeding: happy intestines, healthy breasts, and undetectable viral load. Regarding the first item, signs of intestinal dysbiosis would be indicators for interrupting breastfeeding, such as diarrhea and vomiting, as an imbalance in intestinal flora could create a more favorable environment for infection. The second item addresses pausing breastfeeding in cases of mastitis, as inflammation and recruitment of inflammatory cells could also increase the risk of vertical transmission. Finally, it is necessary for the mother to remain on regular ART and maintain an undetectable viral load for safer breastfeeding (SBP, 2018).

Another issue to address is the potential medication effects of ART on breastfeeding. Issues such as the pharmacodynamics of drugs used in therapy in breast milk, their concentration, and potential effects on the infant’s body are poorly studied. Pharmacovigilance of infant exposure is scarce, with underreporting of adverse effects and potential cases of drug resistance with the use of dolutegravir in postnatal vertical transmission (WHO, 2010; Lehman et al., 2008).

During the study, the need to enhance laboratory analyses with the collection of intrapartum viral load, to be compared with the viral load present in colostrum, was noted. The initial samples were compared with the viral load collected at 34 weeks of gestation, as this analysis is required by the Ministry of Health for performing a vaginal delivery in women living with HIV (WHO, 2016). Monitoring the preconceptional viral status of the woman is also necessary, as it was observed that women with an undetectable preconception status have higher chances of obtaining undetectable colostrum, and women who reach this level during pregnancy may present colostrum with levels below detection, as in our study, below 20 copies per milliliter. Similar results have been observed in the literature, with the hypothesis of CD4+ T cells being stored in reservoirs close to mammary glands, being transmitted through breast milk even after achieving undetectable plasma (WHO, 2010).

Additionally, it is necessary to consider increased expenses due to the need for postnatal monitoring if breastfeeding is offered. As previously mentioned, a monthly surveillance protocol for the maternal-fetal dyad is necessary, increasing the number of tests to be conducted and preventive health measures with information on signs and symptoms of mastitis and gastrointestinal infections in infants

## Data Availability

All data produced in the present work are contained in the manuscript.

## Funding Information

This work was supported by the Ministry of Health of the Government of Brazil. These funding sources had no role in study design or collection, analysis and interpretation of data.

## Competing Interests

The authors declare that this research was conducted in the absence of any commercial or financial relationships that could be implicated as potential conflicts of interest.

## Ethical Statement

This study made no use of human or vertebrate animal subjects and/or tissues. All authors approved the current version of this manuscript and consented to this publication as a preprint on MedRxiv.

## Acknowledgments

(WHO, 2010; Freeman-Romilly et al., 2019; SBP, 2018; Fogel et al., 2013). On the other hand, breastfeeding could reduce the provision of formulas by SUS, reduce the impacts caused by postpartum depression, and future costs related to breast cancer and infant immunological issues, both preventable through breastfeeding, making it important to analyze the new financial balance that would be established with breastfeeding under these conditions.

We would like to thank the staff of the Gaffrée and Guinle University Hospital, Laboratório de Pesquisa Multiusuário 01 (LPM-01) and the Federal University of the State of Rio de Janeiro. We also thank to Laboratório de Pesquisa Multiusuário 04 (LPM-04) and Laboratório Central of Gaffrée and Guinle University Hospital.

## Notes

### Competing Interest Statement

The authors have declared no competing interest.

### Author Declarations

This study was submitted for evaluation by the Research Ethics Committee on human beings at the Gaffree and Guinle University Hospital (HUGG) of the Federal University of the State of Rio de Janeiro (UNIRIO), registered, and approved under the number CAAE: 61885722100005258

